# Do Networks facilitate innovation, learning and sharing? An evaluation of the Quality-of-Care Network (QCN) in Bangladesh, Ethiopia, Malawi, and Uganda

**DOI:** 10.1101/2023.12.05.23299487

**Authors:** Kondwani Mwandira, Seblewengel Lemma, Albert Dube, Kohenour Akter, Asebe Amenu Tufa, Agnes Kyamulabi, Gloria Seruwagi, Catherine Nakidde, Kasonde Mwaba, Nehla Djellouli, Charles Makwenda, Tim Colbourn, Yusra Ribhi Shawar

**Affiliations:** Parent and Child Health Initiative Trust, Lilongwe, Malawi; Department of Disease Control, London School of Hygiene & Tropical Medicine, based in Ethiopia; Perinatal Care Project, Diabetic Association of Bangladesh, Dhaka, Bangladesh; Ethiopian Public Health Institute, Addis Ababa, Ethiopia; Department of Social Work and Social Administration, Makerere University School of Public Health, Kampala, Uganda; Institute for Global Health, University College London, London, UK; Bloomberg School of Public Health, Johns Hopkins University, Baltimore, USA; Paul H. Nitze School of Advanced International Studies, Johns Hopkins University, Washington D.C., USA

**Keywords:** Quality of Care, Quality of Care Network (QCN), Sharing, Learning, Innovations, Maternal Health

## Abstract

The Quality-of-Care Network (QCN), launched by WHO and partners, links global and national actors across several countries to improve maternal and newborn health. We examine if QCN facilitated learning, sharing and innovations within and between network countries.

We evaluated the QCN in Bangladesh, Ethiopia, Malawi and Uganda. We conducted a total 227 qualitative interviews with QCN actors iteratively across a 3-year period from October 2019 to March 2022. We also reviewed all accessible QCN documents.

Sharing and learning opportunities were evident through a number of virtual and in-person platforms such as meetings, conferences, webinars, and websites. Conferences and webinars have been hosted on online resource libraries such as the quality-of-care network website. These provided access to materials on strategies and approaches shared by different countries and actors. Innovations were also evident in some countries like Ethiopia. Innovative software applications aimed at boosting the capacity of service providers were developed and these were adopted by countries such as Bangladesh. Locally, there had been strengthening of learning collaborative meetings, coaching and mentorship. Regular meetings such as Stakeholder Coordination Meetings and Learning Collaborative Sessions provided an opportunity for stakeholders to strategize, as well as share and learn approaches within the network.

The network has promoted coordination among stakeholders. Similar approaches to sharing and learning such as Learning Collaborative Sessions were evident across the study countries. Innovations were not as apparent across countries and most of the learning and sharing approaches were similar to those from broader, pre-existing maternal health approaches, adopted from an era preceding the QCN. There was evidence that the introduction of the QCN improved the functionality and visibility of learning and sharing platforms.

## INTRODUCTION

Many women and babies continue to die from complications in pregnancy and childbirth due to poor quality of and access to peripartum care, especially in low and middle-income countries (LMICs) like Bangladesh, Ethiopia, Malawi and Uganda [1]. To achieve Sustainable Development Goal (SDG) targets for ending preventable maternal and neonatal deaths, there is need for governments and all stakeholders to deliver on the actions called for in global action plans and global strategy [2-4], which require sustained and locally led implementation efforts.

Although in recent years, there have been improvements in rates of institutional delivery across low-income settings, including Malawi (91% in 2015-6 up from 55% in 2000), Bangladesh (37% in 2014 up from 12% in 2004), Uganda (73% in 2016 up from 57% in 2011) and Ethiopia (26% in 2016 up from 10% in 2011) [5, 6]. These improvements in access to care have not been accompanied by improvements in quality, and institutional maternal and neonatal mortality remain very high. For instance, institutional neonatal mortality in Malawi slightly increased from 8.3 in 2017/2018 to 9 for every 1000 live births in both 2018/19, while maternal mortality was at 439 for every 100,000 live births in 2015-16 [7]. While effective interventions exist to treat the main causes of maternal and neonatal deaths in LMICs, effective implementation remains a challenge [8], with contributing factors ranging from insufficient human resources, poor training of health staff, and inadequate infrastructures to shortages in equipment and medications [9], Further, there have been few quality measurements to assess the performance of services and to quantify the gap between reality and expectations in reference to certain standards and guidelines [9, 10].

In response to challenges in the implementation of quality care across many countries, in 2017, World Health Organisation (WHO) and global partners launched ‘*The Network for Improving Quality of Care for Maternal, Newborn and Child Health*’ (QCN) [11]. The aim was to reduce in-facility maternal, neonatal and stillbirth case fatality rates by 50% in five years in nine countries: Bangladesh, Cote d’Ivoire, Ethiopia, Ghana, India, Malawi, Nigeria, Tanzania and Uganda [12]. To achieve this main aim, WHO and partners sought to build a cross-country platform for joint learning around quality improvement implementation approaches based on a shared theory of change (ToC) and shared health outcome goals championed by promotion of partner coordination while emphasising country ownership, leadership and accountability, and shared learning [13].

Learning, one of the strategic objectives in the WHO’s Learning, Accountability, Leadership and Action (LALA) Framework [14], focused on three prominent outcomes namely: (1) data systems development or strengthening to integrate and use quality of care data for improved care (2) development and strengthening of mechanisms to facilitate learning and sharing of knowledge through a learning network and (3) analysis and synthesis of data and practice for an evidence base on quality improvement (Figure 1). These three objective outcomes were developed on the background that individual countries and the global community have much experience of what does and does not work, and each country needs to adapt good ideas to its local context. Therefore, it was presumed that a learning network would allow the transfer of information and knowledge freely between and within all countries interested in improving quality of care.

Drawing on the third objective (learning) of the WHO Learning Accountability Leadership Action (LALA) framework, this paper examines if and how QCN facilitated learning, sharing and innovations within and between network countries. We investigate how the attributes of the network, its operational strategy and performance affect learning, sharing, and innovations among network actors at global and national level.

## METHODS

This analysis was part of a broader multi-country evaluation of QCN [15-18], common methods are reported in our methods supplement for our QCN evaluation collection of papers (S2 Text). Key aspects of the methods in relation to this paper are summarized below.

### Research setting and Population

This research was conducted in Bangladesh, Ethiopia, Malawi, and Uganda; selected case study countries in our broader QCN Evaluation study (S1 Text). We conducted key informant interviews (KII) with selected individuals from the Ministry of Health and stakeholders at the national, district and facility level. To complement our findings, we further conducted non-participatory observations of service provision in selected facilities at the local level. KIIs and observations at the facility level were conducted in four facilities in each country. These four facilities were selected at the onset of the research in 2019. The selected facilities included two high performing and two low performing (Table 1). This performance-based selection was based on maternal and new-born health outcomes which included comparison of maternal and child mortality indicators and other quality of care data (for example those used in national schemes). The QCN survey was extended to all facility stakeholders within selected QCN learning districts in each country: learning facilities across 7 districts in Bangladesh, 6 learning districts in Malawi, 6 districts in Uganda and 8 regions and city administrations in Ethiopia.

### Data used

#### Key Informant Interviews

We conducted semi-structured interviews with selected national and local level network members and key stakeholders in Bangladesh, Ethiopia, Malawi and Uganda between October 2019 and May 2022. At the national level, we interviewed actors from the Ministries of Health (MoH) and key implementation partners. Beyond the national level, we conducted qualitative interviews with key quality improvement and QCN representatives in 4 selected districts in each country (Table 1). We paid particular attention to the perspectives and goals of those carrying out the work of the network, with a specific interest on information pertaining to learning and sharing platforms within the network and pertinent innovations linked to delivery of care in facilities and innovations on delivery of strategic management. Two to four longitudinal iterative rounds were conducted in each country to capture changes and evolution of the network operations, including exploring innovations, learning and sharing and views pertaining to network activities as well as follow-up on emerging findings from the previous rounds. The iterative nature of our study included follow-up of emerging findings from the previous rounds. In total we conducted 227 KII at country level (Table 2).

#### Document review

We reviewed all accessible published and unpublished documents and communications relating to the QCN. These included strategy and management documents, operational plans, directives, formal minutes, and reports (Table 3). We were able to access unpublished documents via WHO and MoH’s QCN contacts. These documents aided understanding of how the QCN is enacted at the global level and the national level, and how the two levels are linked to influence delivery of care at the facility level. We also reviewed online accessible resources including websites and YouTube channel specific to the Quality-of-Care Network.

#### Data Analysis

To address the objective of this paper (explore whether QCN facilitated learning, sharing, and innovation), we analysed the qualitative data (KIIs and documents) for each of the four countries across all rounds. We synthesised and populated country specific qualitative data using 3 main themes (Table 4; sharing, learning and innovations). To understand factors that influence knowledge sharing and learning in the global context across countries, we used a common coding framework developed from an underlying theory adopted from Wu SY et al 2014, “*Knowledge Sharing Among Healthcare Practitioners: Identifying the Psychological and Motivational Facilitating Factors*” (Figure 2) to group data [19].

The theory used in this paper [19] hypothesized that Knowledge Sharing Behaviour (KSB) is a result of Knowledge sharing Intention (KSI), However, the theory suggests that KSI is directly influenced by Psychological Empowerment (path H3e) or indirectly through intermediate predictors such Organization-based psychological Ownership (path H3a), Knowledge-based psychological Ownership (path H3b) and Autonomous Motivation (path H3d). In Summary, Figure 2 indicates relationships between several predictors and outcomes in the theory; ranging from Psychological Empowerment, immediate determinants of Knowledge Sharing Intention (intermediate predictors) to Knowledge Sharing Behaviour. In this paper, we use the theory as a guide to present global results on Psychological Empowerment and Psychological Ownership (organisation-based and knowledge-based) as direct and intermediate predictors that influence KSI.

Psychological empowerment focuses on personal perceptions and empowering experiences and how organizations build a conducive environment for these. Organisation-based psychological ownership can be built via knowledge sharing behaviours, improved teamwork, and communication and coordination, and can facilitate changes in behaviour that can improve practices in quality of care and service delivery, and ultimately clinical outcomes [19, 20].

#### Ethics

All interviews were conducted after obtaining informed written consent from the participants, including separate consent for tape recording. All data is confidential and anonymised. Ethical approval was obtained from the Research Ethics Committee at University College London (3433/003), National Health Sciences Research Committee in Malawi (Protocol number: 19/03/2264), Institutional Review Boards in Bangladesh (BADAS-ERC/EC/19/00274), Ethiopian Public Health Institute in Ethiopia (EPHI-IRB-240-2020) and Uganda (Makerere University School of Public Health Higher Degrees Research Ethics Committee, Ref: 869).

## RESULTS

### Global level

We present global level results systematically under two themes adopted from a learning and sharing theory by Wu et al 2014 [19]. Foremost, we present results on psychological empowerment, being the first main driver of learning and sharing intents, and psychological ownership (Organisation and Knowledge based) being the intermediate predictor of learning and sharing intentions and behaviour [19]. We further explain how these influence learning and sharing across member countries at the global level. We also present a narrative of findings on platforms available globally for learning and knowledge sharing.

### Psychological empowerment

Actors from both the global and national level found QCN to be an important opportunity to facilitate learning and sharing knowledge on quality-of-care practices among countries; allowing countries within the network the ability to learn and adopt working strategies and practices from other countries with similar contextual factors. Further, sharing of strategies was perceived to be a medium to finding common solutions to address causes of maternal and child mortality that are prevalent in LMICs such as sepsis, birth asphyxia and other childbirth complications. QCN’s establishment was largely motivated by the observation that different countries were approaching problems around maternal, newborn and child health in different ways, and that countries could benefit from more information sharing among countries.

> *“Sharing of information…is important for learning. The problem we are dealing with in our country might have been dealt with by some other country. Maybe that other country addressed that problem successfully which can be used to solve our country’s problem. Then, some learning of our country may be helpful to other countries.” **Round1-National-BGD***

One major component of psychological capability for QI in member countries like Bangladesh was adequate training and reporting systems, which enabled improved monitoring, as well as learning from identified deficiencies. QCN-related activities in Bangladesh, particularly the efforts of UNICEF, Save the Children-IHI, and QIS, focused on introducing new data systems and training hospital staff to use them. Global level webinar training for national level actors also acted as a tool for psychological empowerment for in-country actors to spearhead implementation.

### Psychological Ownership - Knowledge based and Organisation Based

The operational design of the QCN fostered network ownership and motivated in-country actors to share and learn. Specifically, QCN’s enactment enabled actors at each level (national and facility) to take the activity implementation lead and champion implementation independently, at the same time ensuring that global and national actors oversee implementation at the national level and the district level or facility level respectively. For instance, in Malawi and Bangladesh, below the QCN coordination team at the national level, local focal persons (District and Facility QI Coordinators) and Quality Improvement Support Teams (QISTs) were selected within their implementation level to oversee the QCN implementation at their own respective implementation level too, imparting a sense of ownership. QIST for example operated at management level and comprised heads of departments (or representatives) and were responsible for overseeing quality improvement at their respective health facilities and reporting the same to a quality focal person within their district and facility. Below the QI focal persons, each MNCH ward was also motivated by having its own Work Improvement Team (WIT), which was responsible for implementation of QI projects at ward level based on the common issues that they face, mostly informed by data over a specified period. The national level stakeholder’s major responsibility was to provide strategic support, while implementation was done by local teams. For instance, in Malawi, the quality improvement plans developed by the WITs outline the QI efforts they undertake based on the needs and gaps identified through the baseline assessments facilitated by the Quality Management Directorate (QMD). This in turn encouraged high participation by QISTs and WITs representatives at collaborative meetings and knowledge sharing events.

> *“The fact that the institutions can identify problems is one result in itself. The development of quality projects, identifying problems, solving those problems and sharing lessons with others is a manifestation in itself. Designing the projects, implementing and reporting for the federal government and the federal government to make them available for others to share experiences is a great manifestation” **Ethiopia Round1-Local”***

### Sharing and Learning Platforms

#### Virtual and in-person meetings for sharing and learning within the QCN

Globally, the network offered several platforms for rapid sharing across its member countries, with the ability to bring high-level professionals from different countries and regions together to discuss key issues relevant to the network. This was centrally organized by WHO and UNICEF.

Countries were brought together through virtual forums like webinars and conference calls. Where feasible, face-to-face meetings were also conducted. Webinars had been conducted since the onset of the QCN in 2017, and they were more prominent from the year 2020 as the onset of the Covid 19 pandemic stalled regular face-to-face meetings between network members; ultimately, proving to be much less costly compared to face-to-face meetings which require huge resources to bring people together.

One prominent example of a virtual meeting was the WHO coordinated meeting called “*integrated approach for implementing MNCH-QOC,"* conducted in August 2020, where Malawi and Uganda along with other member countries were in attendance. During this meeting, experts from the WHO and partners led the mentorship of countries on several issues such as strategies to ensure sustained quality experience of care for clients in facilities. Prominent face-to-face meetings include the March 2019 meeting in Addis Ababa which had French-English bilingual translation services to allow participation of Francophone countries. The meeting provided both plenary and focused small group opportunities for the 11 network countries and 11 observer countries to share their experiences and progress of working towards improved quality of care.

#### Website-based platforms for sharing, learning and accessing QCN resources

Dedicated online websites were also created to serve as a key resource for all network related material [14]. For instance, a QCN dedicated YouTube channel was established in March 2019 where several webinars were posted. As of the 15th of September 2023, the channel had 695 subscribers, 102 videos posted and 29079 views [21]. The webinars posted mainly served as a refresher for the “*WHO MNCH Quality of Care Guidelines*” and a space for countries to share implementation progress.

From the webinar series available on the QCN dedicated website, a few data oriented innovations have been evident. For example, an online application called “Safe Delivery App” [22], which was shared by stakeholders from Ethiopia in 2020. According to the webinar presentation, this application was designed to provide service providers with evidence-based and up-to-date clinical guidelines on most common childbirth procedures through animated instructions and videos. Countries including Bangladesh, Guinea, Ghana and India adopted the customised version of the application. In India, for example, from 2018 to 2021, there have been more than 100,000 downloads of the application; 86,242 active application users employ it as a job aid or self-learning tool. It is used in 698 districts across all 36 states and union territories (UTs) [23].

The QCN dedicated website was designed with multiple functionalities which, among others, would enable country actors to explore practical resources such as all QCN updates and reports. By grouping updates, reports and other webinars by country, the “learn from other countries section” also provided an opportunity for country actors to explore country specific QCN material.

### Country-specific learning, innovation, and sharing

We present country specific results on learning, innovations and sharing at the national and local level. This representation is also in line with the LALA strategic objective number 3: mechanisms to facilitate learning and to share knowledge through a learning network (Figure 1).

### Malawi

At the national level, a QCN Coordination Team was established in 2019. Its operations were coordinated by the Ministry of Health’s Quality Management Directorate (QMD) with a dedicated officer responsible for stakeholder coordination [18]. With funding from partners such as WHO, UNICEF, GIZ and USAID, regular meetings were held by this committee, among others to review and develop strategic documents and brainstorm quality improvement implementation progress and provide strategic planning and technical support to the network. Between 2017 and 2021, the stakeholder’s coordination team met quarterly, and sometimes at non-regular intervals depending on the availability of funding.

> *“In terms of coordination at national level there is a coordination team which is comprised of partners as well as government, which meets almost on monthly basis just to discuss how the whole network in terms of performance and implementation how it is progressing… it meets on monthly basis” **Round1-National-MWI-MoH***

Key outcomes of these coordination meetings included strategic documents such as “Maternal New-born Child and Adolescent Health Quality of Care Roadmap” [24] and “The Malawi MNH Standards” [25]. These and other QCN strategies were shared to learning facilities through Trainings of Trainers (ToTs) led by the national level stakeholders. These ToTs were meant to build local capacity and create ownership and hence ensure sustainability. Those trained were expected to take the lead in teaching their fellows in their facilities. Beyond the national level, learning and sharing was carried out through quarterly collaborative learning sessions at zonal and district levels. Learning facilities came together to share and showcase various QI project approaches and successes being carried out in their respective MNCH wards. Facilities also presented their quantitative data on how they have performed on reducing in-facility child and maternal mortalities. Those doing better in one aspect would share what they were doing to attain such an achievement so that others could learn and adopt strategies. Facilities doing poorly also shared challenges during these sessions to discuss possible solutions.

> *“Ok, in these meetings we normally discuss the indicators, on how we are faring on maternal and neonatal indicators. So, we meet like clusters we group these health facilities in clusters whereby we meet for some hours and each facility present their data on how they have performed in antenatal, labor delivery, postnatal and from there we discuss others learn from other facilities where they are doing better and others facilities which are not doing better, they tell us their problems why they are not doing better that part and we help each other coming up with solutions to address the challenges.” **Round2-National-MWI-MoH***

In some districts, learning sites were brought together to share notes on progress and challenges on a quarterly basis. Beyond these learning sites, partners also supported integrated collaborative sessions between quality-of-care learning sites and the other sites within the district so that the learning sites could lead and showcase what they are doing so that the other facilities could learn.

> *“We made sure was to bring together these learning sites so that we can share notes on a quarterly basis… notes on the progress and challenges and then we also used to support the integrated collaborative learning sites, where we had now the quality-of-care learning sites versus the other sites. So, we could bring all the health centers like at the cluster level, taking into consideration that those which are the learning sites could now lead and showcase what they are doing so that others can learn. That was the key initiative that we were doing………….” **Round3-National-mw-Implementation Partner***

### Bangladesh

Since early 2020, several learning and sharing platforms within the QCN have been evident. These included national webinars for digital trainings for hospital staff, WhatsApp groups between various district staff, virtual group calls between district officials, and in-person meetings between facility level staff. Bangladesh’s internal Quality Improvement system has a long-term focus on information sharing between various districts and facilities. Respondents reported engaging regularly with both national and international channels of communication, including through webinars, poster presentations, and newsletters. These channels of communication proved more useful and prominent as the COVID-19 pandemic progressed in 2020, enabling QCN members to adapt to the restrictions posed by the pandemic.

> *“In pre-COVID period, we had some activities on knowledge sharing, one activity was across the facility learning visit that we say experience sharing visit, so that we did among our QED facilities. Staff of new 4 facilities visited old facilities. So, we did not have to provide with so many trainings there because we found that [facility visit] very effective. They visited those facilities and they replicated those quickly. So that we did before COVID. During COVID, through these virtual platform and webinar, we shared learning and practices with each other.” **Round2-National-BGD-Implementation Partner.***

As of June 2021, one project, which was operating in 86 hospital facilities including some learning sites, introduced quality improvement committees and ward improvement teams in the health facilities. It also established district-level learning networks across these teams for sharing strategies and progress of QI initiatives. As part of its planning, the project explicitly intended to “facilitate cross-learning" within the WHO Quality of Care Network.

Several means were carried out to support knowledge management in Bangladesh. As of November 2021, the National Institute of Preventive and Social Medicine (NIPSOM) was developing its skills and strengthening its systems to support QI efforts in the country. They were developing “QI coaches”, who in turn would provide hands-on support to facilities in using QI methods, building systems to disseminate lessons on successes and failure related to efforts to improve quality of care, and building capacity to manage QoC data. The nature of knowledge transmitted through the QCN network in Bangladesh mainly focused on best practices and relatively organic conversations with national-level leaders from other countries.

> *“Since there was no history of quality improvement activities in one hospital, one partner had to start with basics including introducing staff to QI approaches, human resource management, and mentorship. They supported this through various technical training as well as first-hand learning e.g. PDCA as well as learning sessions” **Round2-National-BGD-Implementation Partner.***

### Uganda

With the arrival of Covid-19 in 2020, online spaces like Zoom and Webinars provided an ideal space for sharing and learning within the QCN in Uganda for national and facility-level actors. Learning sites interfaced via zoom meetings with the national level to share their QCN implementation experiences. Respondents found the zoom meetings useful as they learned what other facilities were doing. For example, in one facility, their success with MNCH management was partly driven by the many resources which they had for referral, unlike the other facilities who did not have some referral resources at the time.

> *“……. Directly you send a coin, and you demand an improvement. Today I have said there is no stethoscope you give me, and you say to get a stethoscope if you don’t want to give me money procure it for me. They should allow me to the make the decision. Like right now all the incubators we have are old ones……." **Round1-National-Uganda***

Online spaces such as WhatsApp groups were also found to be feasible, innovative, and cost saving means for closing the communication gap between national and facility level actors. Virtual spaces provided a similar environment to the face-to-face meetings and trainings at a sustainable cost.

Generally, partners at both the national and district level put together guidelines for QoC to train health workers and support the learning and sharing processes. At local level, health workers continued to learn and sought ideas from other QCN facilities. Departmental Continuous Medical Education was responsible for bringing Antenatal Clinic (ANC) improvements teams together.

Those within facilities would share challenges, identify gaps and then agree on recommendations and action plans, and later, assign a responsible person to make sure the action plans are implemented. There were also collaboration links between the ANC and the maternity teams at the same facility. Maternity heads relayed their duty roster to the ANC so that the ANC would inform them of the person on duty at the right time. For most of the facilities in Uganda, one of the most notable aspects learned by front line healthcare workers was on the usage of oxygen systems. Respondents indicated a huge gap prior to the QCN. With the onset of the QCN, health care workers at one facility were trained in oxygen therapy and case management.

> *“I have noticed that this initiative would work better because many of the health workers didn’t know how to operate oxygen systems, so in line of QCN I would say yes, oxygen therapy and case management using oxygen therapy is one of the of the initiatives that would work better to improve QI” **Round1-National-Uganda***

Training manuals using the Quality-of-Care training packages for MNH as well as simplified QI guide for facilities that were implementing MNH standards were also being developed at the national level. As of 2021, participants indicated plans to have a national level space to gather the implementers of Quality of Care to learn how to make improvements in this area. They also appreciated implementation success because of QCN implementation. While the change in leadership and organisation had facilitated more regular meetings and engagement between partners, this was yet to be fully organised, documented, and synthesised for sharing to be put into practice.

> *“I think that one of the important things that is supposed to show that the network is maturing, is if learning is being effectively generated, shared and used for implementation. So, if you ask me whether that is happening, I will say yes. If you ask me whether that is happening in an organized manner, I will not be very sure about what to tell you.” **Round1-National-Uganda***

### Ethiopia

In Ethiopia, at the national level, notable sharing and learning platforms such as Telegram channels were used for the quality improvement work in the regions where MNCH projects were implemented. QCN specific works were also posted on the same group for the wider MNCH group to appreciate. Regional level meetings and learning collaborative sessions also provided a platform to strengthen sharing and learning.

In regional meetings, stakeholders shared experiences from the implementation of the QCN. During these meetings, learning facilities were given an opportunity to present their QI projects and presenting facilities were advised by experts from the national level on areas to improve and maintain. Rewards and recognition were given to well performing facilities. Nonetheless, facilities that were doing better were on a performance decline due to negligence to sustain substantial QI projects.

> *“… Last year, we were awarded money from the federal government. We couldn’t be awarded that if we were presenting it only at regional level. At a stage where the federal government prepared it was presented and they were satisfied; then we were recognized and awarded. After we returned with the award, and to have a better service, there are some inputs needed” **Round2-Local-Health Center-03***

Collaborative Learning Sessions and coaching were also available at the local level for sharing best quality improvement projects. Usually conducted for a period of three days, the hospitals and health centers in the region presented the best practices in QI projects, providing an opportunity for learning and addressing challenges. Coaching programs, which occurred once or twice a year, also provided an opportunity for learning to the frontline healthcare workers. Although supportive supervisions were conducted as a way of transferring knowledge, these were perceived to be of less value compared to the coaching and mentorship programs.

> *“We used to do supportive supervision based on a checklist but now we have changed that to coaching. Coaching is onsite based on their area of interest” **Round2-Local-Hospital-Eth-04***

Some facilities shared their story on the approaches they employed to decrease ANC 1 to 4 dropout rate. Although this sharing has occurred across facilities, some respondents felt that it was not easy to adopt the strategies to fit in their context because of differences in geographical status and population. As such, although the national level strategic direction could be unified, approaches to quality-of-care improvements may be different from facility to facility. Respondents indicated that adoption of QoC strategies across facilities should be data driven, however, there hasn’t been any evidence to support the feasibility of any intervention or approach before adoption.

> *“…………………. You need to collect data which incorporates process. It should be evidence based. Being evidence based, the data which we have got on learning can fill can correct gaps. So, the network has improved the process of quality improvement. It enhanced commitment; brought learning; mobilized resources. So, I think these are the benefits of networking” **Round2-National-Eth-Implementation Partner***

## DISCUSSION

The emergence of the quality-of-care network contributed to sharing and learning across member countries and their corresponding QCN facilities. Development and strengthening of mechanisms that facilitate knowledge sharing, as well as those related to data system improvement were evident at the global, national and local levels.

The secretariat for the network and stakeholders at the national level led efforts to create knowledge sharing platforms between countries. A number of conferences and webinars were hosted, providing access to materials on strategies and approaches within the network. Strengthening of learning collaborative session meetings, coaching and mentorship were also evident at the national and local level, despite some of these not necessarily specific to the QCN. This serves as evidence of the progress made by the QCN to achieve the learning strategic objective outcome on setting up mechanisms to facilitate learning and to share knowledge through a learning network.

Although most of the national level actors compared to facility level actors have actively been involved in several global level meetings, their attendance has hardly contributed to transforming the day-to-day operations of the QCN in facilities [16]. Resources and approaches learned from other countries have not substantively descended to facility level actors by national level stakeholders. Further, there has not been a substantive number of innovations reported across the network countries. Whilst the network has created systems and contributed to improved circumstances for learning, innovation and sharing for quality improvement, further improvements and investments in individual and organizational capacities are required to effect greater changes [18]. Implementation has also often followed a MNCH holistic approach to quality improvement, not necessarily specific to the network itself. For instance, several participants had challenges distinguishing conferences meant for QCN or quality improvement in general. Further, implementation funding has often been donor driven [15, 16]; this may also pose as a challenge to the sustainability of the network activities [17].

National-level respondents indicated that it was not easy to test ideas learned because the implementation partners responsible for channeling resources had other priority areas. Mentorship, coaching and collaborative learning sessions have been key for individual learning facilities to interface with each other and the national level to share projects and progress being made. Many of these activities were pre-existing before the network. However, no real adoption of approaches learned has been manifested and a lack of funding also affected their implementations, especially for facility specific learning sessions between quality improvement teams. A lack of a coherent evidence generation system to qualify intervention process contributed to lack of evidence to distinguish what qualifies as a working intervention and why the intervention works. Monitoring data for quality improvement was generally found to be a work in progress with many gaps in the availability, accuracy and use of data remaining, as corroborated by our related study on the effectiveness of QCN [16].

This qualitative research investigated whether health networks facilitate innovation, learning and sharing, learning from an evaluation of the Quality-of-Care Network (QCN) in Bangladesh, Ethiopia, Malawi, and Uganda. Firstly, the research methods demonstrate an insightful and contextually rich approach, utilizing KIIs and document reviews to capture the multilevel and multidimensional aspects of the network. The use of qualitative methods aligns well with the exploratory nature of the research topic, allowing for a deep exploratory understanding of learning and sharing mechanisms in the QCN network. Further, our KIIs guides were not rigid across rounds of data collection, we reviewed and revised the topic guides before each round to make sure that pertinent issues from the previous round were explored during the follow up round.

Additionally, the sample selection was purposive, encompassing a diverse range of network actors from various levels of the network. This deliberate sampling strategy enhances the transferability of findings to a broader population for a multilevel network evaluation. As a limitation, the study misses substantive data on innovations at country level within the network hence presenting little evidence on innovation emanating from the network because.

It is unclear how the Psychological Empowerment; the immediate determinants of Knowledge Sharing Intention and Knowledge Sharing Behaviour are clearly linked in a multilevel health network and whether these linkages are significant. Future research could focus on this to advance understanding of how it influences sharing, learning and innovation in a health-related network.

## CONCLUSION

Health networks are important to fostering learning and sharing. The network promoted coordination among stakeholders, and several sharing platforms and meetings were conducted to equip countries with implementation updates and facility QoC approaches. Similar approaches across the study countries to sharing and learning, for example, Learning Collaborative Sessions, were evident. However, innovations were not as apparent across countries due to the broader pre-existing maternal health approach adopted from an era preceding the QCN by countries towards its implementation. There was evidence however that the introduction of the QCN improved the functionality of learning and sharing platforms.

## Data Availability

All data is derived from interviews conducted with stakeholders and document reviews in each country. Careful consideration has been taken to ensure anonymity of the data in the submitted manuscript. Sharing a de-identified data set would jeopardize the conditions of informed consent. Data requests can be made to each of the Ethical Review Committees who approved this research and must be accompanied by a detailed explanation of how the data will be used in line with the ethical approvals received. Contact information for each committee are as follows: University College London Research Ethics Committee (3433/003) phone: +44 20 7679 8717, BADAS Ethical Review Committee (ref: BADAS-ERC/EC/19/00274) phone: +880 5861664150, Ethiopian Public Health Institute Institutional Review Board (ref: EPHI-IRB-240-2020) phone: +251 11 2133499, National Health Sciences Research Committee in Malawi (ref: 19/03/2264) phone: +265 789 400, and Makerere University Institutional Review Board (ref: Protocol 869) phone: +256 414 705500.

## ACKNOWLEGEMENTS

We thank all respondents and stakeholders for their time and contributions toward making this work possible. The QCN Evaluation Group is: Nehla Djellouli, Kasonde Mwaba, Callie Daniels-Howell, Tim Colbourn (UCL Institute for Global Health, UK), Kohenour Akter, Fatama Khatun, Mithun Sarker, Abdul Kuddus, Kishwar Azad (BADAS-PCP Bangladesh), Kondwani Mwandira, Albert Dube, Gladson Monjeza, Rachel Magaleta, Zabvuta Moffolo, Charles Makwenda (Parent and Child Health Initiative, Malawi), Mary Kinney, Fidele Mukinda (independent researchers, South Africa), Mike English (Oxford University), Yusra Shawar, Will Payne, Jeremy Shiffman (Johns Hopkins University, USA), Kathy Lubowa, Agnes Kyamulabi, Hilda Namakula, Gloria Seruwagi (Makerere University, Uganda), Anene Tesfa, Asebe Amenu, Theodros Getachew, Geremew Gonfa (Ethiopia Public Health Institute, Ethiopia), Seblewengel Lemma, Tanya Marchant (LSHTM, UK).

